# Psychological preparedness for pandemic (COVID-19) management: Perceptions of nurses and nursing students in India

**DOI:** 10.1101/2020.09.24.20201301

**Authors:** Sailaxmi Gandhi, Maya Sahu, G Radhakrishnan, Prasanthi Nattala, Paulomi M. Sudhir, Rathi Balachandran

**Affiliations:** Department of Nursing, National Institute of Mental Health & Neuro Sciences (NIMHANS), Bengaluru, Karnataka, India; Department of Clinical Psychology, National Institute of Mental Health & Neuro Sciences (NIMHANS), Bengaluru, Karnataka, India; Nursing Division, Ministry of Health and Family Welfare, Government of India, New Delhi, India

**Keywords:** Psychological Preparedness, Pandemic Management, COVID-19, Nurses, Nursing Students

## Abstract

**Introduction:** The growing COVID-19 pandemic has posed a great threat to millions of people worldwide. Nurses and nursing students are an important group of health professionals who are most likely to face many challenges in this unprecedented scenario. The present study aimed at exploring the perception of nurses and nursing students regarding psychological preparedness for the pandemic (COVID-19) management.

**Materials & Methods:** The study employed a quantitative cross-sectional online survey research design. Purposive sampling was used with an attempt to represent the entire nurses (i.e. nursing officers, nurse administrators and nursing teachers) and nursing students group of India. The survey link was shared to their email ID and they were invited to participate in the study. Data were collected using Psychological Preparedness for Disaster Threat Scale (PPDTS)-Modified, General Self Efficacy (GSE) Scale, Optimism Scale and Brief Resilient Coping Scale (BRS). Totally 685 responses were received and 676 forms were completed which were analyzed using SPSS software (version 24).

**Results:** The mean age of the subjects was 31.72 (SD=9.58) years. Around 20% of the subjects previously had some kind of psychological training and 4% of the subjects had taken care of persons with COVID-19. Findings revealed that mean score for PPDTS, GSE, BRCS and Optimism was 73.44 (SD=10.82, 33.19 (SD=5.23), 16.79 (SD=2.73) and 9.61 (SD=2.26) respectively indicating that the subjects had moderate level of psychological preparedness, self-efficacy and resilience but higher level of optimism. Psychological preparedness, self-efficacy, optimism and resilience were positively correlated to each other. Self-efficacy, optimism, and resilience emerged as predictors of psychological preparedness.

**Conclusion:** The findings suggested that self-efficacy, optimism and resilience can be considered as predictors for psychological preparedness in pandemic management. Appropriate training could influence self-efficacy while programs addressing resilience and coping may strengthen psychological preparedness which can help in further management of ongoing pandemic.

## Introduction

Disasters or natural hazards are a significant source of psychological distress. Reactions to the trauma associated with natural disasters, hazards are varied, including adverse effects on the psychological well-being of individuals exposed to these events. There is a significantly greater burden of mental health conditions in countries affected by disasters and trauma-related experiences. [1,2]

The growing pandemic of Coronavirus (COVID-19) caused by a newly discovered coronavirus, has posed a great threat to millions of people worldwide. [3] Although there have been various pandemics and disease outbreaks such as plague outbreaks, Cholera pandemic, Spanish Flu, Asian flu, SARS, MERS, Ebola, and Zika, the socio-economic disruption and psychological impact caused by COVID -19 has been enormous. As with most medical/biological disasters, health professionals are the frontline workers who deliver essential health care services and hence are the first point of contact.

The COVID-19 outbreak is a unique and unprecedented scenario for many health care workers, particularly if they have not been involved in similar situations [4]. Worries about patient care, adequacy of protection, long working hours, inadequate access to basic needs, stigma due to the risk of infecting others and separation from families can lead to severe psychological distress among them Further, as the pandemic progresses, they are becoming more and more engaged in COVID-19 care. [5] Nurses (i.e. nursing officers, nurse administrators and nursing teachers) and nursing students are an important group of health professionals who are at risk of facing several similar challenges pertaining to deliver of health care.

Preparedness is an important variable in the context of disasters. The key influences on preparedness include risk perception; preparedness perceptions such as outcome expectancy; self-efficacy; collective efficacy; previous experience; perceived responsibility; responsibility for others; coping style; and resource issues. [6] Psychological preparedness is an important aspect of various types of preparedness. Reser & Morrissey (2009) describe psychological preparedness as an *“intra-individual and a psychological state of awareness, anticipation, and readiness capacity to anticipate and manage one’s psychological response in an emergency situation”*. It is an important resource for health providers during times of natural/ biological disasters. It is determined by psychological factors such as self-efficacy, optimism, state-trait anxiety and resilience. [7]

While there has been much research on psychological preparedness in the context of natural disasters, there is a paucity of work on psychological preparedness, particularly amongst nurses, in the context of biological disasters. There is little research on the identification of elements that constitute disaster-related psychological preparedness in nurses. [1] Hence, it is important to understand their perception of psychological preparedness for this pandemic. Understanding how they perceive their psychological preparedness will shed light on the available strength and resources allowing the possibility of developing a shared set of objectives for a better plan of interventions and a more sensitive pandemic management approach. Hence, the present study aimed at exploring nurses’ and nursing students’ perception of psychological preparedness for the pandemic (COVID – 19) management. The objectives were: 1) To assess the psychological preparedness for pandemic management, self-efficacy, optimism and resilient coping of nurses and nursing students in India. 2) To find out the relationship between psychological preparedness, self-efficacy, optimism and resilient coping. 3) To identify the association between socio-demographic variables and the study variables.

## Materials & Methods

### Research approach and design

This study employed a quantitative cross-sectional online survey research design.

### Subjects

Nurses and nursing students from central, state Government and private hospitals were invited to participate in the study.

### Sampling and setting

Purposive sampling was used with an attempt to represent the entire nurses (i.e. nursing officers, nurse administrators and nursing teachers) and nursing students’ group of India. The subjects were identified from a pool of college/universities/registered names in various professional organizations. Totally 685 responses were received and 676 forms were completed which were considered for analysis.

### Sampling criteria

Registered nurses or nursing students studying diploma, graduate, postgraduate or PhD Nursing and both genders were included in the study. The nurses or nursing students who did not have access to internet and hence could not participate in the online survey.

### Procedure

After identifying the potential subjects, the survey link of Google form was shared to their email ID including the tools for the study. Those who agreed to take part filled up some baseline information such as age, education, details of current work. After this, the next two sections contained questions on overall preparedness for events and also factors that influence it. Completing these tools took approximately 15 to 20 minutes of their time. Data confidentiality and anonymity were ensured from the researchers’ end.

### Method of data collection

An online survey link was shared to the subjects and willing subjects were invited to fill the form containing the questions.

#### Measures

Data were collected using the following tools:

##### Part-I: Demographic Variables

A detailed schedule to document demographics was prepared by the researcher and validated by experts. It included information on demographics along with information regarding work and experience in the area of disaster management as well as specific experience in working with persons with COVID-19.

##### Part-II: Psychological Preparedness for Disaster Threat Scale (PPDTS) - Modified

The PPDTS is a scale with 26 items on a 4-point Likert-type scale. Psychometric properties of the PPDTS demonstrate that the scale is a valid and reliable measure of psychological preparedness [2]. The scale showed excellent internal consistency, with a Cronbach’s alpha of 0.93. For the present study, the tool was modified after obtaining permission from the authors. Cronbach’s alpha for in this sample was 0.94.

##### Part-III: General Self Efficacy (GSE) [8]

It contains 10 questions on a 4-point scale (1-not at all true to 4 - exactly true). The tool has adequate psychometric properties and the internal reliability of this scale ranges from 0.76 and 0.90. [8] Cronbach’s alpha for the present study sample was 0.90. The tool has good construct validity.

##### Part-IV: Personality Predispositions

a. Optimism: According to Carver & Scheier, (2014), dispositional optimism is a cognitive construct that includes expectancies regarding outcomes of future events. The Life Orientation Test, revised version (LOT-R), scale quantifies optimism [10] using a 3 item scale. Three items measure optimism on a 4-point Likert scale: 0 = strongly disagree, 1 = disagree, 2 = neutral, 3 = agree, and 4 = strongly agree. Cronbach’s alpha for the present study sample was 0.71.
b. Resilience and Coping - Brief Resilient Coping Scale (BRS): A brief measure in resilient coping was administered in the present sample. The BRS is a 4 item measure and participants rate these four statements from 1 (does not describe me at all to 5 (describes me very well) Higher scores indicate greater resilient coping. A high score between 17 and 20 – indicates that the person is a highly resilient coper, and a low score – between 4 and 13 – suggests that the person is a low resilient coper. Reliability (Cronbach’s alpha) of the tool for the present study sample was 0.83.

### Ethical consideration

The research protocol was reviewed and approved by the Institute Ethics Committee. An online survey link was shared to the subjects with electronic consent form where subjects were explained about the aims and objectives of the study and requested to participate in the study. Informed consent was obtained from the willing participants and confidentiality was maintained. All the participants were given the freedom to withdraw from the study whenever they wanted.

### Data analysis

The data were analyzed using statistical software (SPSS 24 version) and p < 0.05 was considered as the level of significance. Descriptive statistics such as mean (SD)/ frequency (percentage) were used to describe the socio-demographic variables as well as scores of study variables. Independent sample t-test was used to assess the differences of scores in binary variables and One-way ANOVA with post hoc tests was conducted to find the association between the Chi-Square test was used to find the differences of scores of study variables with categorical demographic variables. Correlation between the study variables and few demographic variables was assessed by using Pearson correlation coefficient. Multiple linear regression was used to predict the variables of PPDTS. Thematic analysis was done for the open ended questions in the demographic data sheet.

## Results

The final sample comprised of 676 nurses and nursing students. The mean age of the subjects was 31.72±9.58 years. Majority of them were female (77.8%), married (55.3%), completed M.Sc. Nursing (54%), were teaching staff/ faculty (46.6) and from non-Mental Health Speciality (73.8) (Table 1).

**Table 1:** Socio-demographic variables n=676.

| **Variables** | **Categories** | **Frequency (%)** |
| --- | --- | --- |
| Age |  | 31.72 ± 9.58<sup>#</sup> |
| Years of work experience |  | 5 (1.5, 12)<sup>α</sup> |
| Gender | Males | 150 (22.2) |
|  | Females | 526 (77.8) |
| Marital status | Never married | 291 (43) |
|  | Married | 374 (55.3) |
|  | Separated/widowed | 11 (1.6) |
| Education level | GNM | 41 (6.1) |
|  | B.Sc. | 231 (34.2) |
|  | M.Sc. | 365 (54) |
|  | Ph.D. | 39 (5.8) |
| Job title | Student | 251 (37.1) |
|  | Teaching staff/ Faculty | 315 (46.6) |
|  | Nursing Officer/Administrator | 110 (16.3) |
| Practice speciality | Mental Health Nursing | 177 (26.2) |
|  | Non-Mental Health Nursing | 499 (73.8) |
| Current place of work | Clinic/Hospital | 137 (20.3) |
|  | Educational Institute | 529 (78.3) |
|  | Community health facility | 10 (1.5) |
| Type of institute | Central Govt | 144 (21.3) |
|  | State Govt | 98 (14.5) |
|  | Private | 410 (60.7) |
|  | NGO | 24 (3.6) |

<sup>#</sup>Mean±SD, <sup>α</sup>Median (Q1, Q3)

Previous experience in disaster management was reported by 11% of the sample. Types of disaster management indicated included Tsunami, flood relief, cyclone, fire burns, trauma care and as student volunteers. Around 20% of the subjects previously had some kind of psychological training and 67.5 of them had once, 15.4 had twice and others had undergone training for more than two times. Around 4% of the subjects had taken care of patients who were COVID-19 positive (Table 2).

**Table 2:** Experience related to psychological training in disaster management n=676.

| **Variables** | **Categories** | **Frequency (%)** |
| --- | --- | --- |
| Previous experience in disaster management | Yes | 76 (11.2) |
|  | No | 600 (88.8) |
| Types of disaster management | Tsunami | 13 (17.1) |
|  | Flood relief | 18 (23.7) |
|  | Cyclone/storm/Landslide | 4 (5.3) |
|  | Fire/Burns | 4 (5.3) |
|  | Trauma care | 8 (10.5) |
|  | Student volunteer | 5 (6.6) |
|  | Others | 24 (31.6) |
| Previous psychological training | Yes | 121 (17.9) |
|  | No | 555 (82.1) |
| Nature of training | Mock drill/ basic course | 26 (21.5) |
|  | Continuing Nursing Education (Workshops/conference/lectures/hospital training) | 33 (27.3) |
|  | Disaster management (earth quake/ fire/ trauma) | 54 (44.6) |
|  | Psychological first aid/crisis intervention/emotional counseling | 5 (4.1) |
|  | First response training/Basic Life Support (BLS) | 3 (2.5) |
| Taken care of COVID patients | Yes | 26 (3.8) |
|  | No | 650 (96.2) |

Findings revealed that mean score for PPDTS, GSE, BRCS and Optimism was 73.44±10.82, 33.19±5.23, 16.79±2.73 and 9.61±2.26 respectively indicating that the subjects had moderate level of psychological preparedness, self-efficacy and resilience but higher level of optimism (Table 3).

**Table 3:** Scores of the study variables.

| **Variables** | **Score range** | **Mean ± SD** |
| --- | --- | --- |
| **Psychological preparedness** | 26-104 | $73.44 \pm 10.82$ |
| **Self-efficacy** | 10-40 | $33.19 \pm 5.23$ |
| **Optimism** | 0-12 | $9.61 \pm 2.26$ |
| **Resilience** | 5-20 | $16.79 \pm 2.73$ |

Psychological preparedness, self-efficacy, optimism and resilience were positively correlated to each other. Significant positive correlation was also found between these variables, age and work experience (Table 4).

**Table 4:** Correlation between the study variables.

| Variables | Psychological preparedness | Self-efficacy | Optimism | Resilience | Age | Work experience |
| --- | --- | --- | --- | --- | --- | --- |
| **Psychological preparedness** |  |  |  |  |  |  |
| **Self-efficacy** | 0.781** |  |  |  |  |  |
| **Optimism** | 0.443** | 0.468** |  |  |  |  |
| **Resilience** | .624** | .697** | .549** |  |  |  |
| **Age** | .287** | .231** | .154** | 0.196** |  |  |
| **Work experience** | .281** | .213** | 0.127** | 0.168** | 0.918** |  |

\*\* p<0.01

There was a significant association between psychological preparedness, self-efficacy, optimism, resilience and few demographic variables. The PPDTS score was significantly higher among the nursing faculty and administrators than the students (F=25.46, p<0.001), among those who were from mental health nursing (MHN) speciality than non-MHN (t=3.95, p=0.001), who had previous training in disaster management than who did not have (t=3.26, p=0.001) and the nurses who were working in private institute than those who were working in central or state govt. institute (F=4.21, p=0.006).

Similarly, GSE score was higher among the nursing faculty and administrators than the students (F=15.15, p<0.001), among those who were from MHN speciality than non-MHN (t=2.59, p=0.010), and the nurses who were working in private institute than those who were working in institutes under the central government (F=5.10, p=0.002).

Further, optimism and BRCS scores were higher among the nursing faculty than the students. BRCS score was significantly higher among those who were from MHN speciality than non-MHN (t=2.03, p=0.043) and optimism score was also significantly higher among the nurses who were working in private institute than those who were working in institutes under the central government (F=2.19, p=0.08) (Table 5).

**Table 5:**
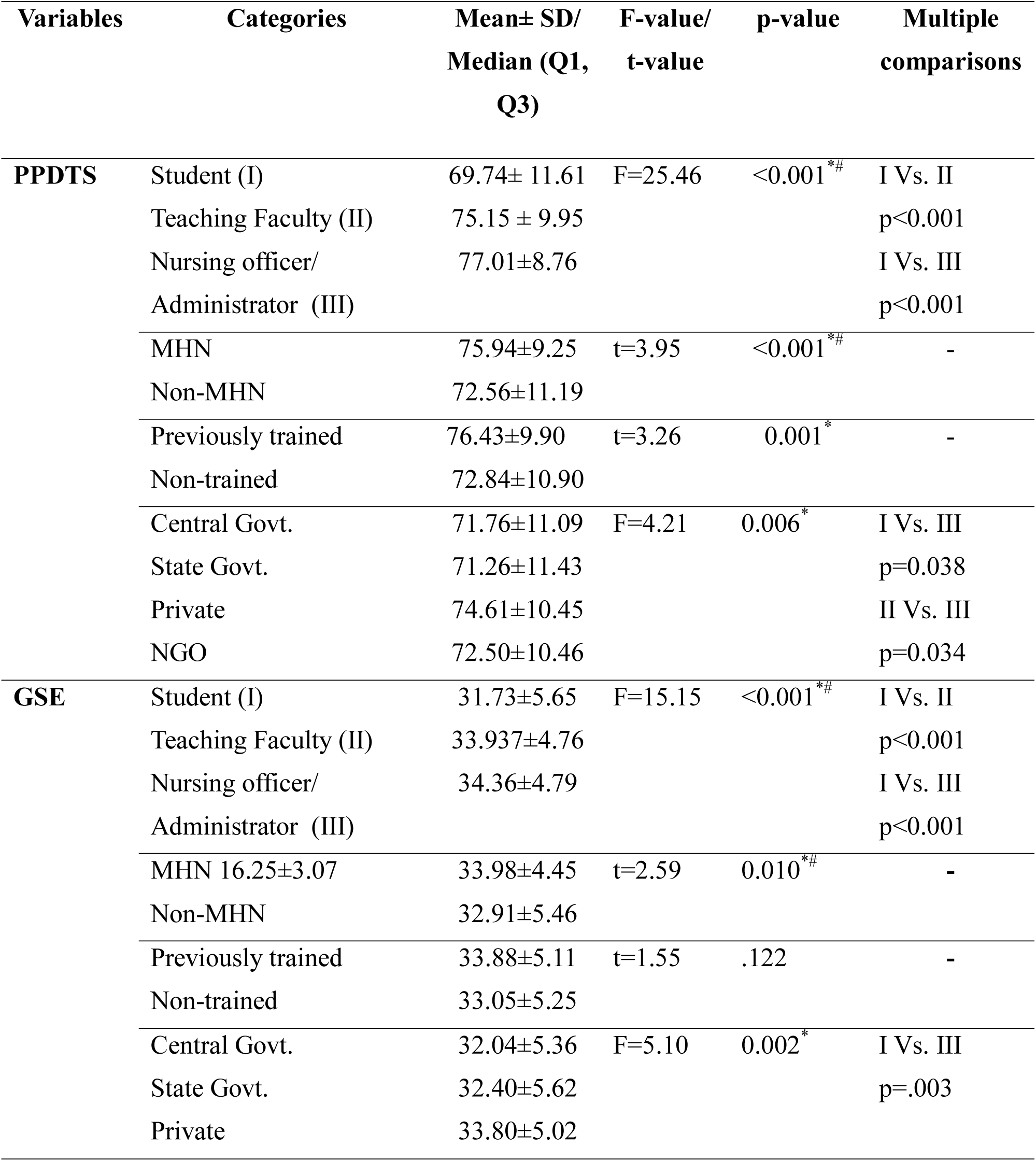
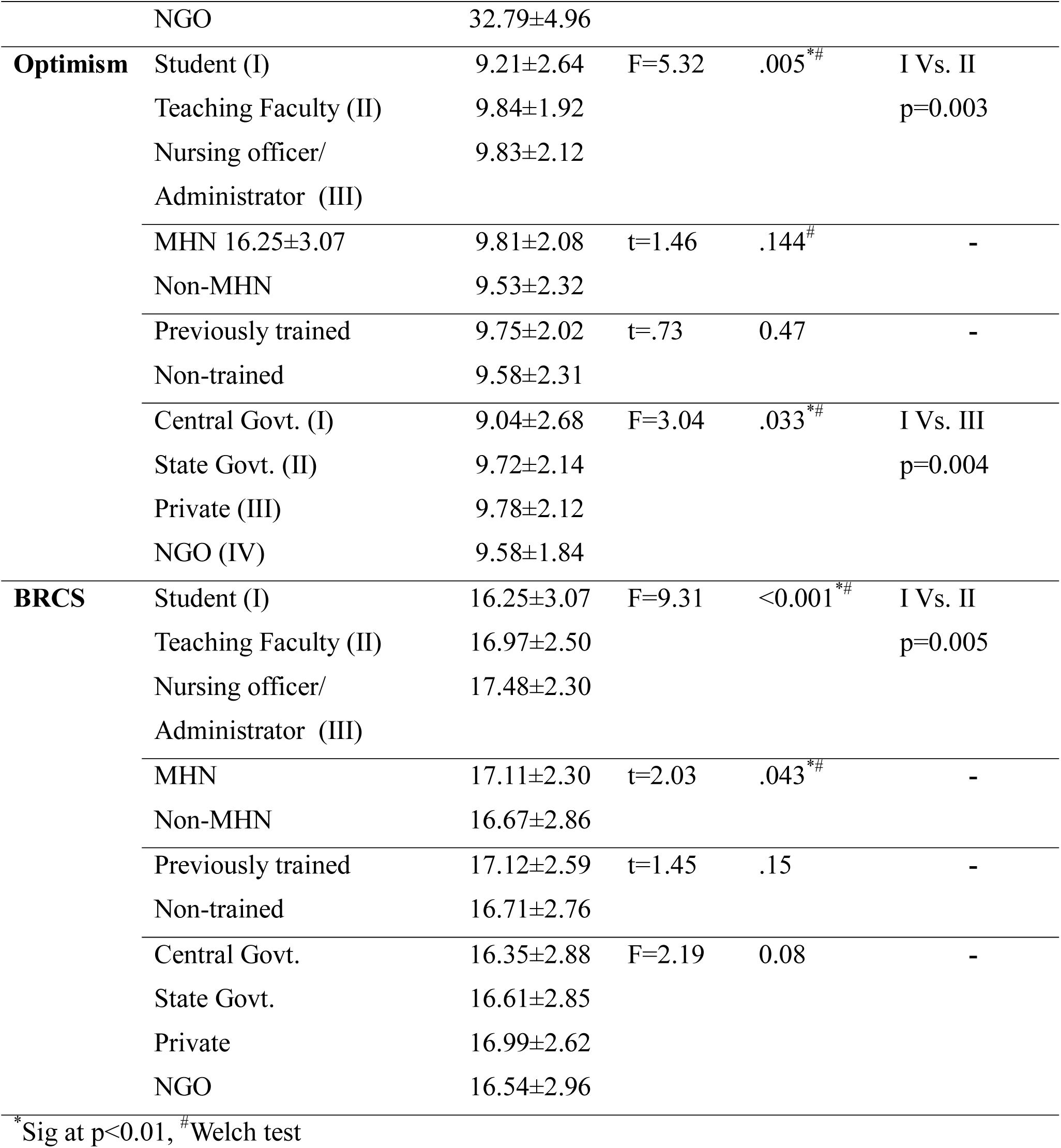
Association/Comparison among psychological preparedness, self-efficacy, optimism and resilience with selected demographic variables.

Self-efficacy, optimism, and resilience emerged as predictors of psychological preparedness. Multiple linear regression analysis found that that self-efficacy, optimism and resilience were able to explain 62% variance in psychological preparedness (Table 6).

**Table 6:** Predictors of psychological preparedness:

| Predictors | B(SE) | Std Beta | t-value | p-value | R square |
| --- | --- | --- | --- | --- | --- |
| Intercept | 16.64 (1.8 ) |  | 9.448 | <0.001 |  |
| GSE_TOTAL | 1.37 (0.07 ) | 0.662 | 19.886 | <0.001 | 0.623 |
| OPT_TOTAL | 0.3 (0.14 ) | .062 | 2.171 | .030 |  |
| BRCS_TOTAL | 0.51 (0.14 ) | 0.128 | 3.644 | <0.001 |  |

### Qualitative findings

Thematic analysis of the open-ended questions posited two major themes: *positive experiences* and *suggestions given*. The subjects who took care of COVID 19 positive patients had shared their positive experiences and gave few suggestions that would be useful for others (Table 7).

**Table 7:** Themes and sub-themes emerged from the data n=26.

| Themes & Sub-themes | Codes |
| --- | --- |
| **I. Positive experiences** |  |
| *1. Gained confidence* | • Capability in disaster management<br>• Management of critical situations<br>• Felt motivated and encouraged |
| *2. Satisfaction of care giving* | • Recovery of patients including elderly and younger ones<br>• Availability of resources<br>• Serving humanity and nation<br>• Admiration and appraisal from patient and caregivers<br>• Recognition from supervisor |
| **II. Suggestions given** | • Stick to the protocol<br>• Proper precaution<br>• Avoid being panicky<br>• Provide support<br>• Correct use of learnt skill<br>• Provide correct information<br>• Adequate self-confidence<br>• Positive thinking<br>• Motivational talks |

### I. Positive experiences

#### 1. Gained confidence

All the nurses who have shared their positives experiences mentioned that they became more confident after caring for persons with COVID positive. As stated by few of them,

> *“It became my learning experience and learned how to work in disaster management*.*”*
>
> *“I have learnt how to deal with COVID positive patient. Motivating to the patient is very important. Encourage the patient for taking medication and maintenance of social distances is important*.*”*

#### 2. Satisfaction of care giving

Though there were challenges at times, most of the participants felt that the satisfaction of providing care to the persons with COVID positive was very gratifying, as expressed by some of them,

> *“When I was on COVID duty, we treated a 76-year-old male patient whose test report became negative after 3 positive reports. Similarly, a small 4-year-old baby had a negative report after 3 positive reports. These gave me a lot of satisfaction*.*”*
>
> *“The satisfaction that the patient recovered completely…. The testimonial and the feedback that we receive from the patient’s attenders are really good. Though we’re working in the Suit, the words of encouragement that we get from our superiors are worth the work*.*”*
>
> *“When both patients got discharged and our 14 days shift was over with no staff infected, we were so happy at that time”*
>
> *“Self-Satisfaction in life-saving process and in service to the needy…”*

### II. Suggestions given

The participants urged others to follow protocol, take precautions and promote mental health of patients as well as self. As verbalized by few of them,

> *“Always try to communicate with the patient, reassure them. Donning and doffing of PPE is very important. Most of the time we get infected because of improper handling of PPE, and last but not the least don’t be panic*.*”*
>
> *“Support each other and be mentally strong…”*
>
> *“Consider every patient as a positive patient. Secondly, be well prepared for the procedures to reduce the time taken at patient’s bedside*.*”*
>
> *“Adhere to protocol… do frequent hand washing, maintain social distance and be confident*.*”*
>
> *Please don’t worry about the infection risk. Adhere to social distancing norm at home, working environment and all possible environment. Please educate at least one person in your vicinity about COVID 19. There by you’ll not only educate them, but you’ll spread positive awareness*.*”*

## Discussion

COVID -19 pandemic has placed healthcare professionals across the world in an unprecedented situation, having to make impossible decisions and work under extreme pressures which may be difficult for some to handle the situation properly [11]. The present study explored nurses’ and nursing students’ perception about psychological preparedness for pandemic (COVID - 19) management. Our findings are consistent with previous findings.

Only around 11% of the subjects in the present study had previous experience in disaster management and around 20% had some kind of psychological training. Of this, continuing nursing education or basic mock drill was comparatively more than formal disaster management training. However, this experience of either facing disaster earlier or having training might influence the psychological preparedness of the nurses or nursing students. While few studies documented that previous exposure to disasters may effect risk perception and the degree of preparedness, others reported no influence of individual’s previous experience of disasters with preparedness. [2,12]

Around 4% of the participants had cared for patients who had tested positive for COVID-19. This pandemic is quite different from the previous disasters which had caused havoc in various ways. Their experience of working with COVID positive patients may encourage or motivate others to come in the frontline or who are already working with fear, anxiety or stress due to a lot of undue challenges.

Participants in the present study reported moderate level of psychological preparedness, self-efficacy and resilience but higher level of optimism. It was also found that these variables were positively correlated to each other. A study conducted by Said et al., (2020) among 88 nurses who had at least one occasion of experience in responding to a disaster, found that around 50% of the respondents perceived a high level of PPTDS with a high mean self-efficacy.[1] Similar to the current study findings, this study also documented that psychological preparedness was positively correlated with self-efficacy, optimism, self-esteem, older age, more years of experience and more times responding to disasters. PPDTS had a strong positive correlation with self-efficacy and a high positive correlation with resilience suggesting that presence of self-efficacy [13] and resilience helps individual handle difficult situation and conversely, the higher the psychological preparedness, the more the perceived self-efficacy or resilience. Further, high level of optimism among the present study subjects may contribute to a positive relationship with adaptive coping that might enhance their cognitive and emotional functioning when they face challenges [14] or may have less psychological distress and greater resilience to post-disaster psychological sequels. [15]

Self-efficacy, optimism, and resilience emerged as predictors of psychological preparedness. Multiple linear regression analysis found that self-efficacy, optimism and resilience were able to explain 62% variance in psychological preparedness indicating that individuals with higher self-efficacy, optimism and resilience are more likely to be better psychologically prepared for management of pandemic. Said et al., (2020) also found that self-efficacy and self-esteem predicted 53% variance of psychological preparedness.[1] In another study, small-scale computer-based simulation exercises were found to be effective in improving head emergency nurses’ general self-efficacy and disaster management skills.[13] Thus the findings also highlight the need to work on improving these areas among health care professionals to equip them to handle difficult situations properly.

Psychological preparedness, self-efficacy, resilience and optimism were higher among the nursing faculty and administrators than the students. It is quite natural for the younger students to lack in these areas which might improve with age and work experience. According to Ministry of Health & Family Welfare (MoHFW), Govt. of India, if the need arises, students can be involved in handling crisis as per their level of skills and training. For example, 1^st^ and 2^nd^ year undergraduate nursing students can be utilized to take care of Non-COVID-19 patients whereas 3^rd^ and 4^th^ year undergraduate nursing students in the caring for mild to moderate COVID-19 patients. Post graduate and graduate (Post basic) nursing students being registered Nursing Officers can be utilized to take care of severe COVID-19 patients. [16] However, providing measures like psychological first-aid and counselling may help them be better prepared particularly when the student nurses have to shoulder such additional responsibilities in an emergency.

Perceived psychological preparedness and resilient coping scores were also higher among those who were from MHN speciality indicating that being in Mental Health Nursing field may be advantageous when psychological preparedness is considered. However, this does not indicate that non-MHN professionals are not in a position to handle difficult situations properly. Rather, providing psychological training might help them improve their well-being as well as functioning. Psychological preparedness before and during a pandemic may enable individuals to identify their feelings and to manage these cognitive and emotional responses [7] so that they can better focus on management of pandemic.

Participants’ positive experience of taking care of persons with COVID positive and the suggestions given would help others gain some ideas and avoid errors in their practices. Though there are challenges while caring for the persons who are COVID positive, the satisfaction gained at the end of the day motivates them to provide care even with more passion. Similar findings were echoed in an earlier study on psychological experience of nurses caring for COVID-19 patients from China. The study mentioned 4 emerged themes: 1) presence of negative emotions in early stage consisting of fatigue, discomfort, and helplessness 2) self-coping styles including psychological and life adjustment, altruistic acts, team support, and rational cognition. 3) growth under pressure, which included increased affection and gratefulness, development of professional responsibility, and self-reflection and 4) emergence of positive emotions simultaneously with negative emotions [17]. Reiteration on adherence to protocol and maintaining hygiene is the reflection of precautionary measures to be taken by the health care personnel as well as the common public [4].

## Strengths & limitations

The main strengths of the present study were large sample size and inclusion of nurses and nursing students from all over India which may be representative of the country and hence increases the generalizability of the findings. The present study also included nurses who had experience of taking care of persons with COVID 19 positive (although less n=26) and documented their experiences which is very first of its kind in the Indian setting. Further, using open-ended questions in addition to standardized tools enhanced the richness of the data. The present study was not without certain limitations such as cross-sectional survey design. The findings should be interpreted cautiously as it does not provide cause and effect relationship rather provides the possibilities of the direction which needs to be studied in future. Further, the possibility of social desirability bias due to self-reported questionnaire could not be avoided. Despite the limitations, the present study provides important findings on psychological preparedness in an important section of health care professionals i.e. nurses and nursing students. Aspects of psychological preparedness studied here may help in planning further for pandemic management.

## Conclusions

The current study aimed at exploring psychological preparedness among nursing students and nurses during COVID 19 pandemic. The results highlighted the importance of considering psychological preparedness in the planning and management of current pandemic and training for nurses as well as nursing students. Findings suggested that self-efficacy, optimism and resilience can be considered as predictors for psychological preparedness in pandemic management. Further, managing one’s mental health and psychosocial well-being during this time is as important as managing one’s physical health. Appropriate training could influence self-efficacy while programs addressing resilience and coping may strengthen psychological preparedness which can help in further management of ongoing pandemic.

## Data Availability

Data underlying the study cannot be made publicly available due to ethical concerns
regarding participant privacy, as imposed by the Institute Ethics Committee of the
National Institute of Mental Health and Neuro Sciences. Data are available from the
corresponding author (contact) for researchers who meet
the criteria for access to confidential data.

## Acknowledgement

The authors would like to thank to all the subjects for participating in the study.

